# Aspirin-Containing Antiplatelet Therapy and In-Hospital Gout Flares in Acute Ischemic Stroke Patients with Gout and Hyperuricemia: A Retrospective New-User Cohort Study

**DOI:** 10.64898/2026.10.23.26366257

**Authors:** Aiwen Ma, Yu Zhang, Xiaolin Xu

## Abstract

**Background:** Antiplatelet selection is challenging in patients with acute ischemic stroke (AIS), gout, and hyperuricemia. We compared the risk of in-hospital gout flares between aspirin-containing regimens and clopidogrel monotherapy, and examined subsequent aspirin discontinuation during dual antiplatelet therapy (DAPT).

**Methods:** A retrospective new-user cohort study was conducted based on electronic medical records (2019–2025). Patients were categorized into aspirin-containing therapy (aspirin monotherapy or DAPT with aspirin plus clopidogrel) and clopidogrel monotherapy groups. The primary analysis used IPTW-adjusted logistic regression. Interaction tests assessed effect modification and a Sankey diagram visualized DAPT treatment transition.

**Results:** Among 105,890 screened patients, 457 were enrolled (234 aspirin-containing therapy, 223 clopidogrel monotherapy). In-hospital gout flares occurred in 187 (40.9%). Aspirin-containing regimens were associated with a higher flare risk (OR = 1.57, 95% CI 1.07 – 2.31, *P* = 0.022). Results were consistent in sensitivity analyses. Higher SUA (OR 1.20 per 1-mg/dL increase, 95% CI 1.03–1.40; *P* = 0.023) and baseline mRS ≥3 (OR 2.13, 95% CI 1.42–3.20; *P* < 0.001) were independently associated with in-hospital gout flares. No significant interactions were observed between antiplatelet regimens and SUA, mRS or urate-lowering therapy (ULT). Among DAPT recipients, aspirin discontinuation in patients with gout flares was 65.2%, 66.7%, and 56.0% in 2019–2021, 2022–2023, and 2024–2025, respectively, versus 47.8%, 28.6%, and 11.9% in those without flares.

**Conclusions:** In AIS patients with gout and hyperuricemia, initiating aspirin-containing regimens was associated with a higher risk of in-hospital gout flares compared with clopidogrel monotherapy. Gout flares during DAPT were accompanied by frequent aspirin discontinuation, which may limit completion of guideline-recommended DAPT. Prospective studies are needed.

## 1 Introduction

Ischemic stroke is the most common stroke subtype, accounting for over 7.8 million new cases annually worldwide(GBD 2021 Stroke Risk Factor Collaborators 2024). Gouty arthritis, the most common inflammatory arthritis in adults, is a frequent comorbidity among stroke survivors(Dehlin et al. 2020). Among patients with acute ischemic stroke (AIS), both hospitalization and a history of gout independently increase the risk of in-hospital acute gout flares(Dubreuil et al. 2013; Lin et al. 2009), which can cause severe local joint inflammation, prolong hospital stay(Lin et al. 2009), and potentially contribute to long-term functional decline and disability(“Quality of Life and Disability in Patients with Treatment-Failure Gout - PubMed,” n.d.).

Aspirin-containing dual antiplatelet therapy (DAPT) is widely recommended in guidelines as part of early standard care for acute ischemic stroke to reduce early recurrent stroke. Common indications include: (1) acute minor ischemic stroke (National Institutes of Health Stroke Scale [NIHSS] ≤3) or high-risk transient ischemic attack (TIA), for which a 21-day course of aspirin plus clopidogrel is supported(Prabhakaran et al. 2026); (2) minor stroke (NIHSS ≤5) or high-risk TIA, for which a 30-day course of aspirin plus ticagrelor is an alternative(Dawson et al. 2021; Johnston et al. 2020); and (3) symptomatic intracranial atherosclerotic stenosis (ICAS), for which guidelines recommend a course of up to 90 days of aspirin plus clopidogrel(Psychogios et al. 2022). However, low-dose aspirin is theoretically linked to elevated uric acid(Caspi et al. 2000; Segal et al. 2003). In clinical practice, it remains uncertain whether aspirin increases the risk of recurrent gout flares in AIS patients with gout and hyperuricemia. In addition, when such a flare occurs during the acute hospitalization phase, clinicians may discontinue aspirin and revert to clopidogrel monotherapy. This leaves patients without dual therapy at the time of greatest risk for recurrent ischemic events.

Real-world studies on the association between aspirin, serum uric acid (SUA) metabolism, and gout risk have yielded inconsistent conclusions(Caspi et al. 2000; Segal et al. 2003; Li et al. 2021; Zhang et al. 2020; Jiang et al. 2026; Tan et al. 2024; Panagiotopoulos et al. 2025). Most previous studies were not conducted in patients with AIS(Caspi et al. 2000; Segal et al. 2003; Li et al. 2021; Zhang et al. 2020), and recent studies on stroke–gout associations have included relatively few gout events(Tan et al. 2024; Jiang et al. 2026; Panagiotopoulos et al. 2025). Moreover, whether such gout-related changes in antiplatelet therapy is a frequent and clinically relevant remains unknown. Against this background, we conducted a real-world retrospective cohort study to compare the incidence of in-hospital gout flares between aspirin-containing antiplatelet regimens and clopidogrel monotherapy in a cohort of patients with AIS and concomitant gout/hyperuricemia. We further explored the clinical and laboratory factors associated with gout recurrence and subsequent unplanned aspirin discontinuation. By characterizing this gout–antiplatelet therapeutic conflict, we aimed to inform personalized antiplatelet management in this high-risk stroke population.

## 2 Methods

This was a retrospective new-user cohort study. Data were extracted from the electronic medical record database of Tianjin Huanhu Hospital, China, covering the period from 1 January 2019 to 31 December 2025. The study protocol was approved by the Institutional Review Board of Tianjin Huanhu Hospital. Reporting of this study conforms to the Strengthening the Reporting of Observational Studies in Epidemiology (STROBE) guidelines.

### 2.1 Study Population

The study cohort included adult patients admitted with AIS who had hyperuricemia and a documented medical history of gout. AIS was defined as newly diagnosed cerebral infarction confirmed by brain magnetic resonance imaging or computed tomography. Gout was diagnosed according to the 2015 American College of Rheumatology/European Alliance of Associations for Rheumatology classification criteria(Neogi et al. 2015). Hyperuricemia was defined as a fasting serum uric acid level >7 mg/dL (420 µmol/L) (Dalbeth et al. 2021). New users of antiplatelet therapy were defined as patients who had no prior history of antiplatelet use or had discontinued antiplatelet therapy for more than one month before admission, and subsequently initiated antiplatelet therapy within 7 days after admission for the index stroke.

Patients meeting any of the following criteria were excluded: (1) Those receiving antithrombotic agents other than aspirin or clopidogrel (e.g., ticagrelor); (2) Those with a documented gout flare within one month prior to the index date; (3) Those receiving urate-lowering therapy within one month prior to the index date; (4) Those administered medications affecting uric acid metabolism during hospitalization, except benzbromarone, including other urate-lowering agents (e.g., allopurinol, febuxostat), diuretics (e.g., loop diuretics, spironolactone), and antihypertensive drugs (e.g., irbesartan, losartan potassium); (5) Those with severe comorbidities, including renal failure (estimated glomerular filtration rate [eGFR] ≤30 mL/min/1.73 m^2^), Child-Pugh class C liver failure, large cerebral infarction requiring mannitol treatment, or a modified Rankin Scale (mRS) score of 5 or higher.

### 2.2 Exposure Definition and Data Collection

The primary exposure was defined as initiation of one of two antiplatelet groups. The aspirin group included patients on low-dose aspirin monotherapy (100 mg/day or 200 mg/day) or DAPT (aspirin 100 mg/day plus clopidogrel 75 mg/day), with an optional 300 mg clopidogrel loading dose on the first day. The clopidogrel group received clopidogrel monotherapy (75 mg/day). Exposure groups were assigned based on inpatient prescription records, and patients entered the cohort upon hospital admission.

Among patients receiving DAPT, we specifically assessed whether aspirin was discontinued at discharge, defined as the absence of aspirin in the discharge prescription despite its use in the admission DAPT regimen. All DAPT patients were categorized by the primary documented indication for DAPT into three groups: (1) minor stroke, defined as acute ischemic stroke with a baseline NIHSS score ≤5 and symptom onset within 24 hours; (2) ICAS defined as symptomatic 50–99% stenosis of a major intracranial artery confirmed by computed tomography angiography (CTA) or magnetic resonance angiography (MRA); and (3) other indications, including but not limited to progressive stroke, medullary infarction, or other physician-documented conditions requiring DAPT. If multiple indications coexisted, patients were assigned according to a prespecified hierarchy: ICAS > minor stroke > other.

The baseline covariates included sex, age, hypertension, diabetes mellitus (DM), drinking status, mRS score, serum creatinine (SCr), SUA, blood lipids (triglycerides [TG] and low-density lipoprotein cholesterol [LDL-C]), neutrophil count, lymphocyte count, and use of urate-lowering therapy (ULT) with benzbromarone 25 or 50 mg/day. Calculation of eGFR was based on the Chronic Kidney Disease (CKD) Epidemiology Collaboration formula. Neutrophil-to-lymphocyte ratio (NLR) was calculated as neutrophil count divided by lymphocyte count.

### 2.3 Outcome Definition

The primary outcome was incident gout flare, defined as a documented gout flare event in clinical records combined with a prescription for anti-gout treatment. Follow-up started at cohort entry and lasted until the first gout flare or hospital discharge. Length of stay (LOS) was collected.

### 2.4 Statistical Analysis

Missing data were handled using multiple imputation with 5 imputed datasets (chained equations)(White et al. 2011). The missing rates of key variables were as follows: TG 1.97%, LDL-C 1.75%, NLR 3.50%, and eGFR 1.31%. Exposure and baseline covariates except sex, were included in the imputation model. To address confounding, stabilized inverse probability of treatment weightings (IPTWs) were generated, and extreme weights were trimmed at the 1st and 99th percentiles. Covariate balance before and after weighting was assessed using absolute standardized mean differences (SMD). We also verified propensity score overlap and examined the distribution of trimmed weights for stability.

For the primary analysis, a weighted logistic regression model was fitted within each imputed dataset using the generated IPTWs. The estimates were pooled using Rubin’s rules. Odds ratios (ORs) with 95% confidence intervals (CIs) and two-sided P-values were calculated, with clopidogrel monotherapy as the reference group.

Sensitivity analyses were performed to evaluate the robustness of our findings: (1) Overlap weighting as an alternative propensity score approach; (2) An unweighted multivariable logistic regression model, with forest plots generated for visual presentation.

Interaction tests were performed to assess whether the effect of antiplatelet regimens on in-hospital gout flares differed across prespecified baseline factors, including SUA level (per 1 mg/dL increase), baseline mRS score (≥ 3 vs. < 3), and use of ULT with benzbromarone (yes vs. no).

A Sankey diagram was constructed to visualize the distribution and transitions of DAPT patients. Four ordered strata were used: (1) calendar year of admission, (2) primary indication for DAPT, (3) in-hospital gout flare, and (4) aspirin discontinuation at discharge.

Categorical variables were presented as counts and percentages, and compared using the chi-square (χ^2^) test or Fisher’s exact test. Non-normally distributed continuous data were reported as median (interquartile range, IQR), and analyzed via the Mann–Whitney U test. All statistical tests were two-tailed, and a P value < 0.05 was defined as statistically significant. All analyses were performed using R software version 4.5 (2026.01.1 + 403).

## 3 Results

### 3.1 Characteristics of the Study Population

A total of 105,890 patients with AIS were initially screened for eligibility, among whom 1,081 had a history of gout and were new users of antiplatelet agents. After applying exclusion criteria, 457 patients were ultimately included in the analysis. Of these, 234 were assigned to the aspirin group (76 receiving aspirin monotherapy and 158 receiving DAPT), and 223 to the clopidogrel group. The patient screening flowchart is presented in **Fig 1**. Patients in the clopidogrel group tended to be older than those in the aspirin group (median age: 62.0 vs. 59.0 years, *P* < 0.001). No other statistically significant differences were observed between the two groups for the remaining baseline variables. IPTW was applied based on all baseline covariates except sex, owing to the extremely low number of female participants in the cohort. Baseline characteristics of the two groups before and after weighting are shown in **Table 1**. Prior to weighting, 4 of the 11 covariates exhibited an SMD > 0.1. After weighting, all SMDs were below the prespecified threshold of 0.1. The propensity score overlap assumption was satisfied (**Supplementary Fig 1**). Distribution of trimmed weights remained stable (**Supplementary Fig 2**).

**Table 1.** Observed and weighted baseline characteristics of the study population stratified by antiplatelet regimens.

| Characteristics | Unweighted |  |  |  | Weighted |  |  |
| --- | --- | --- | --- | --- | --- | --- | --- |
|  | Aspirin group | Clopidogrel group | Unweighted SMD | P value | Aspirin group | Clopidogrel group | SMD with IPTW |
| Age, years | 59.0 (49.0, 66.0) | 62.00 (54.0, 69.0) | 0.357 | <0.001 | 60.0 (51.0, 68.0) | 60.00 (52.0, 68.0) | 0.008 |
| Sex |  |  | NA | 0.625 |  |  | NA |
| Female, n (%) | 1.0 (0.4) | 6.0 (2.7) |  |  | 1.0 (0.4) | 6.0 (2.7) |  |
| Male, n (%) | 233.0 (99.6) | 217.0 (97.3) |  |  | 233.0 (99.6) | 217.0 (97.3) |  |
| Drinking, n (%) | 152.0 (65.0) | 135.0 (60.5) | 0.091 | 0.379 | 149.3 (63.8) | 141.4 (63.4) | 0.007 |
| HP, n (%) | 190.0 (81.2) | 177.0 (79.4) | 0.046 | 0.709 | 185.8 (79.4) | 177.3 (79.5) | 0.004 |
| DM, n (%) | 44.0 (18.8) | 46.0 (20.6) | 0.046 | 0.709 | 44.7 (19.1) | 42.6 (19.1) | 0.004 |
| mRs (score) |  |  | 0.066 | 0.545 |  |  | 0.009 |
| 0-2, n (%) | 132.0 (56.4) | 133.0 (59.6) |  |  | 136.4 (58.3) | 130.9 (58.7) |  |
| 3-4, n (%) | 102.0 (43.6) | 90.0 (40.4) |  |  | 97.6 (41.7) | 92.1 (41.3) |  |
| SUA, mg/dL | 8.66 (7.85, 9.72) | 8.57 (7.77, 9.89) | 0.015 | 0.971 | 8.67 (7.83, 9.77) | 8.57 (7.77, 9.90) | 0.003 |
| TG, mmol/L | 1.96 (1.33, 2.85) | 1.75 (1.29, 2.40) | 0.114 | 0.070 | 1.81 (1.31, 2.65) | 1.80 (1.31, 2.43) | 0.002 |
| LDL-C, mmol/L | 3.10 (2.55, 3.66) | 3.01 (2.40, 3.73) | 0.052 | 0.460 | 3.06 (2.52, 3.65) | 3.01 (2.43, 3.75) | 0.009 |
| eGFR, mL/min/1.73m<sup>2</sup> | 84.08 (65.15, 102.87) | 77.48 (66.39, 95.47) | 0.110 | 0.126 | 82.90 (64.16, 101.03) | 79.54 (66.60, 98.15) | 0.002 |
| NLR | 2.59 (1.98, 3.69) | 2.87 (2.12, 4.09) | 0.104 | 0.152 | 2.72 (2.01, 3.73) | 2.78 (2.03, 3.87) | 0.003 |
| ULT | 52.0 (22.2) | 47.0 (21.1) | 0.028 | 0.854 | 52.7 (22.5) | 50.4 (22.6) | 0.004 |

Notes: values are presented as median (Q1, Q3) or n (%). DAPT, dual antiplatelet therapy; SMD, stabilized mean difference; IPTW, inverse probability of treatment weighting; NA, not applicable; HP, hypertension; DM, diabetes mellitus; mRS, modified Rankin Scale; SUA, serum uric acid; TG, triglycerides; LDL-C, low-density lipoprotein cholesterol; eGFR, estimated glomerular filtration rate; CKD, chronic kidney disease; ULT, urate-lowering therapy; NLR, neutrophil-to-lymphocyte ratio.

**Fig 1.**
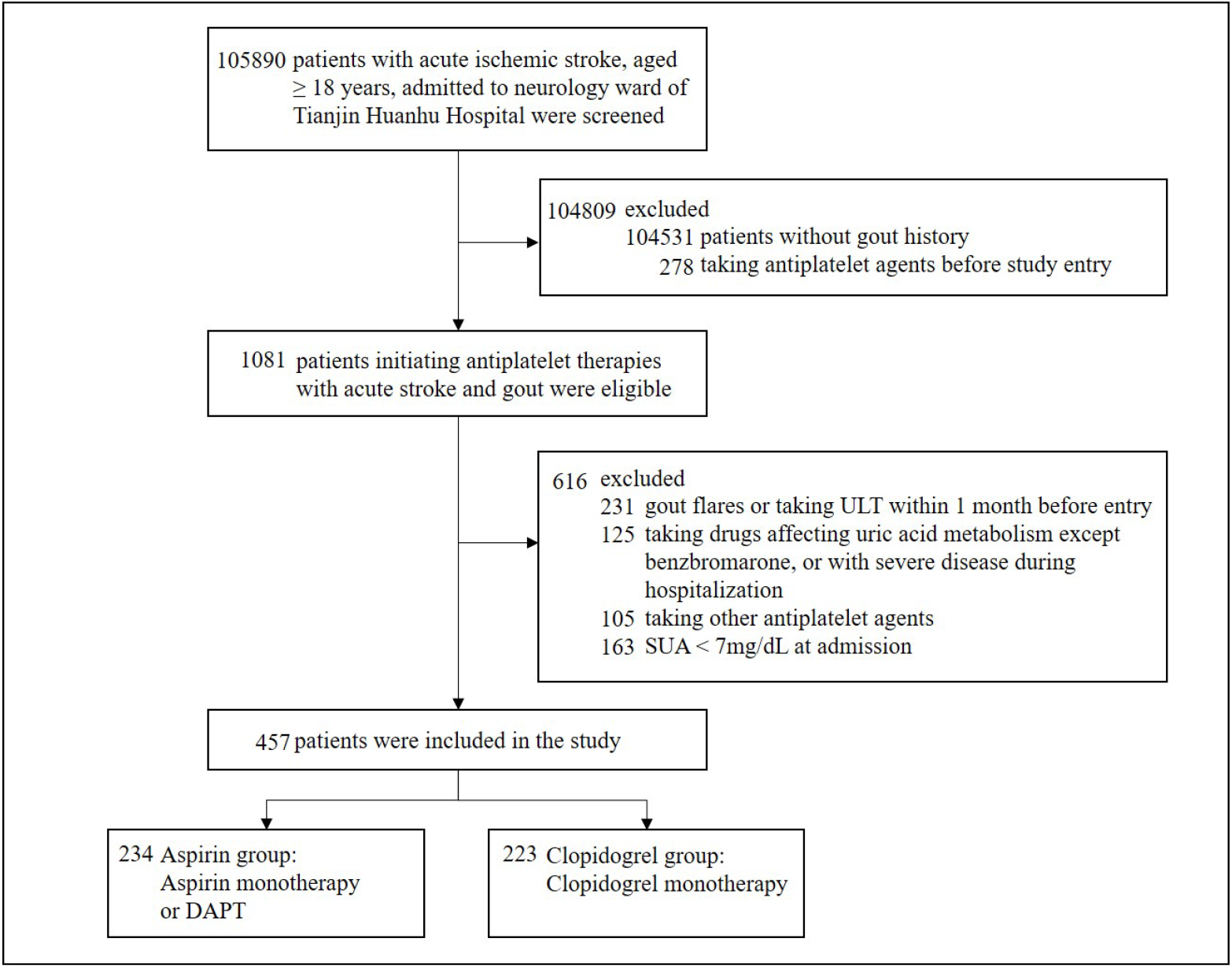
Study flow chart. ULT, urate-lowering therapy; SUA, serum uric acid; DAPT, dual antiplatelet therapy

### 3.2 Primary Outcome

Overall, 187 of 457 patients (40.9%) experienced gout flares, including 109 of 234 (46.6%) in the aspirin group and 78 of 223 (35.0%) in the clopidogrel group. As shown in **Table 2**, after multiple imputation and IPTW-weighted logistic regression, aspirin was associated with a higher risk of gout flare compared with clopidogrel (OR 1.57, 95% CI 1.07–2.31, *P* = 0.022).

**Table 2.**
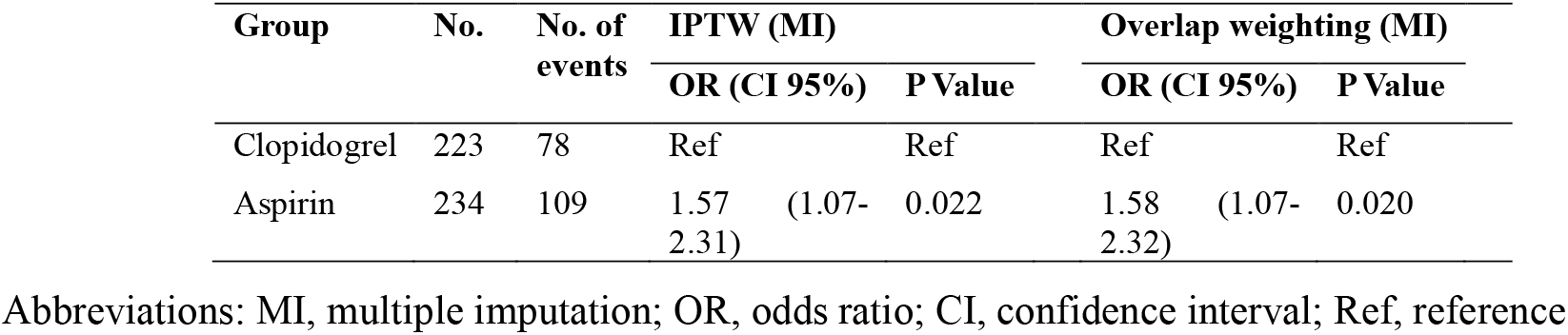
Association of antiplatelet regimens with gout flares under different analytical methods.

### 3.3 Sensitivity Analysis

To evaluate the robustness of the weighting model, we performed sensitivity analyses, both of which yielded results consistent with the primary analysis. First, an overlap-weighted model reproduced the main findings (**Table 2**). Second, in an unweighted multivariable logistic regression model that adjusted for all baseline covariates except sex, aspirin use was independently associated with gout recurrence (OR 1.65, 95% CI 1.10–2.46, *P* = 0.015). A modified Rankin Scale score ≥ 3 (OR 2.13, 95% CI 1.42–3.20, *P* < 0.001) and serum uric acid level (OR 1.20, 95% CI 1.03–1.40, *P* = 0.023) were also independent predictors (**Fig 2**).

**Fig 2.**
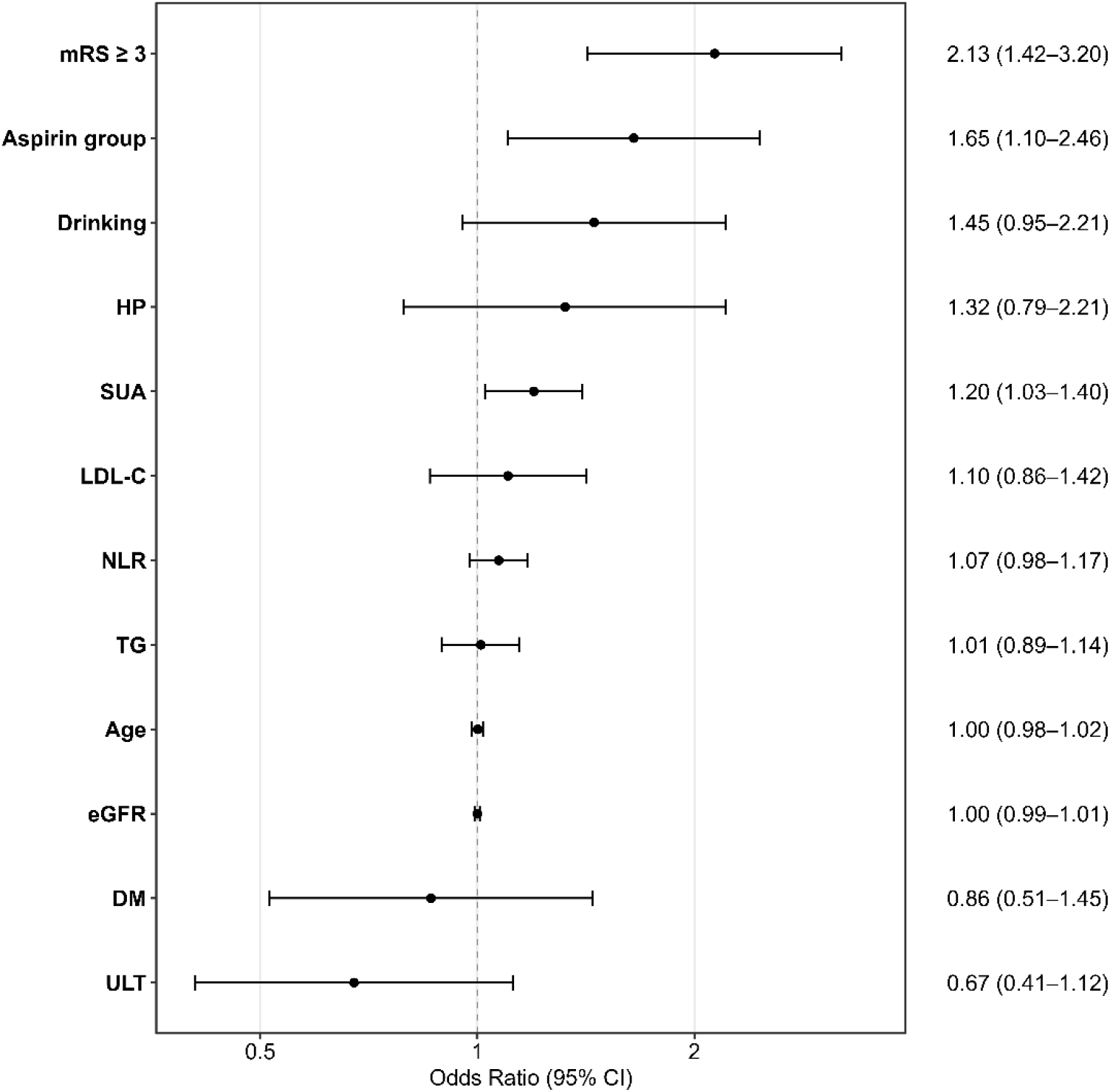
Forest plot displaying aspirin-containing antiplatelet regimens versus clopidogrel monotherapy with adjusted odds ratios (ORs) and 95% confidence intervals (CIs) for in-hospital gout flares. mRS, modified Rankin Scale; HP, hypertension; LDL-C, low-density lipoprotein cholesterol; NLR, neutrophil-to-lymphocyte ratio; TG, triglycerides; eGFR, estimated glomerular filtration rate; DM, diabetes mellitus

### 3.4 Interaction Analysis

IPTW-weighted logistic regression revealed no significant interaction between antiplatelet therapy and ULT, mRS, or SUA (all interaction *P* > 0.05; **Table 3**), suggesting that none of these variables modified the association between antiplatelet strategy and in-hospital gout flares.

**Table 3.** Interaction analysis between antiplatelet regimens and risk factors regarding in-hospital gout flares.

| Interaction term | OR | 95% CI | P Value |
| --- | --- | --- | --- |
| antiplatelet × SUA | 0.81 | 0.60 – 1.09 | 0.170 |
| antiplatelet × mRS | 0.97 | 0.43 – 2.18 | 0.934 |
| antiplatelet × ULT | 0.46 | 0.17 – 1.20 | 0.110 |

Notes: IPTW-weighted logistic regression was applied for interaction analysis.

### 3.5 Distribution of DAPT and Aspirin Discontinuation

In aspirin-treated subgroup, a total of 109 gout flares occurred: 37 of 76 (48.7%) on aspirin monotherapy and 72 of 158 (45.6%) on DAPT. The difference was not statistically significant (*P* = 0.781) after multivariable adjustment for ULT, mRS score, age, SUA, eGFR, and NLR. Treatment adjustments at discharge occurred irrespective of gout flare events: aspirin monotherapy may be converted to clopidogrel monotherapy, and DAPT may be de-escalated by discontinuing aspirin with clopidogrel maintained.

Among all patients, the proportion of DAPT prescription increased gradually across three time periods: 25.1% in 2019–2021, 31.7% in 2022–2023, and 50.8% in 2024–2025. The distribution of DAPT indications differed across periods. The proportion of DAPT prescribed for “other indications” declined from 39.1% in 2019–2021 to 15.0% in 2024–2025, whereas the proportions prescribed for “mild stroke” and “ICAS” increased from 21.7% to 35.8% and from 39.1% to 49.3%, respectively.

Stratified by in-hospital gout flare status, aspirin discontinuation rates among patients initiated on DAPT showed a gradual declining trend over the study period in both groups. Specifically, the discontinuation rate in the flare group remained relatively high and changed only modestly, from 65.2% in 2019–2021 to 66.7% in 2022–2023 and 56.0% in 2024–2025. In contrast, patients without in-hospital gout flare exhibited a marked decline, with discontinuation rates falling from 47.8% in 2019– 2021 to 28.6% in 2022–2023 and further to 11.9% in 2024–2025.

A Sankey diagram (**Fig 3**) illustrates the distribution and transitions of DAPT recipients, and full stratified data are available in **Table 4**.

**Table 4.** Discontinuation of aspirin in patients with DAPT, stratified by year, in-hospital gout flare status, and indication.

| **Gout flare status** | **DAPT indication** | **2019–2021** | **2022–2023** | **2024–2025** |
| --- | --- | --- | --- | --- |
| **Flare** | Mild stroke | 3/5 (60.0) | 8/10 (80.0) | 3/8 (37.5) |
|  | ICAS | 3/7 (42.9) | 4/10 (40.0) | 11/15 (73.3) |
|  | Other | 9/11 (81.8) | 4/4 (100) | 0/2 (0) |
|  | **Total** | **15/23 (65.2)** | **16/24 (66.7)** | **14/25 (56.0)** |
| **No flare** | Mild stroke | 2/5 (40.0) | 4/9 (44.4) | 1/16 (6.3) |
|  | ICAS | 4/11 (36.4) | 0/6 (0) | 1/18 (5.6) |
|  | Other | 5/7 (71.4) | 2/6 (33.3) | 3/8 (37.5) |
|  | **Total** | **11/23 (47.8)** | **6/21 (28.6)** | **5/42 (11.9)** |

DAPT, dual antiplatelet therapy; TIA, transient ischemic attack; ICAS, intracranial atherosclerotic stenosis. Data are presented as n/N (%). N denotes the number of patients in each stratum

**Fig 3.**
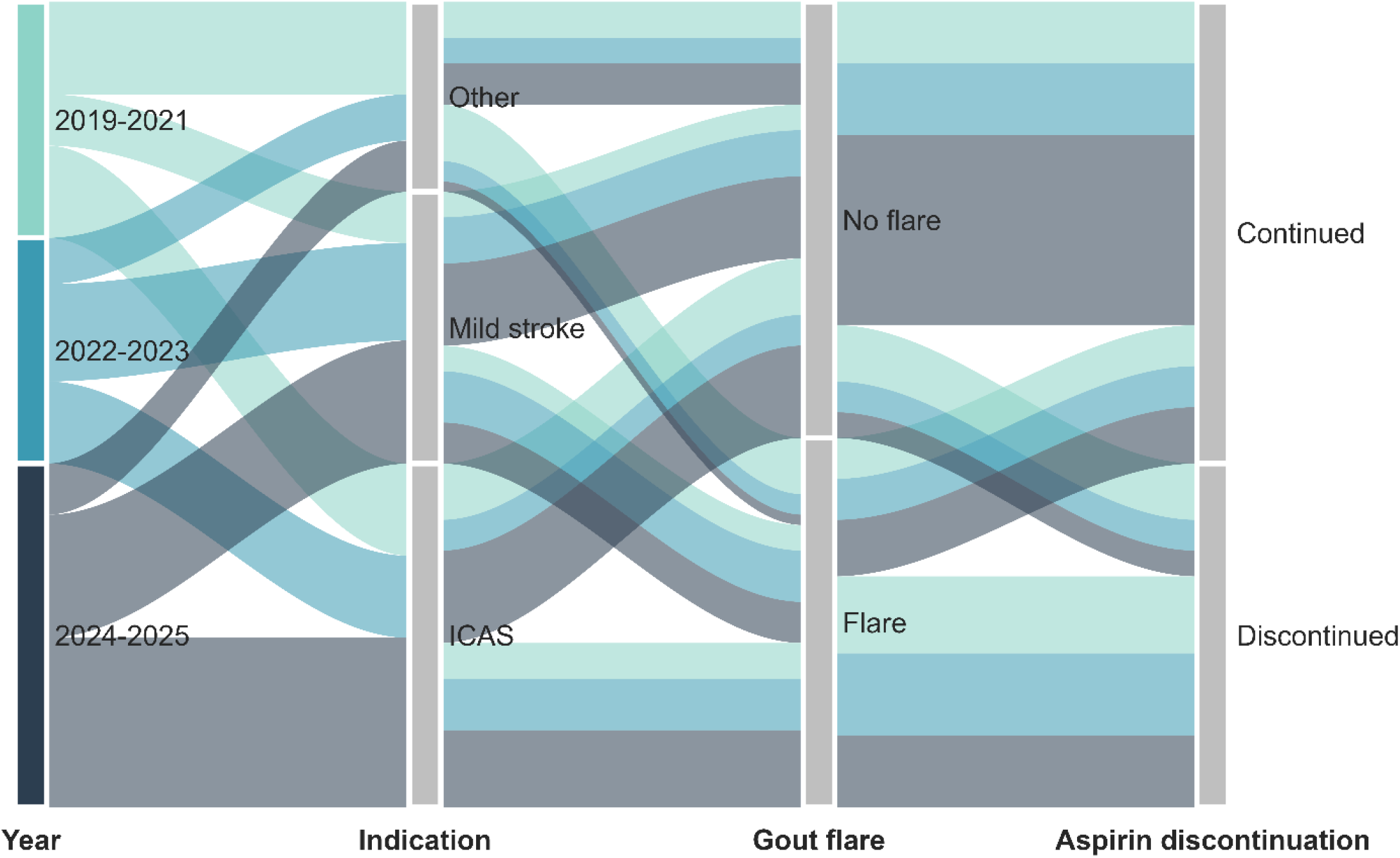
Sankey diagram showing transitions of patients initiated on dual antiplatelet therapy during admission, stratified by study period, DAPT indication, in-hospital gout flare status, and aspirin discontinuation. TIA, transient ischaemic attack; ICAS, intracranial atherosclerotic stenosis

## 4 Discussion

In this retrospective cohort of AIS patients with gout and hyperuricemia, aspirin-containing antiplatelet regimens were independently associated with a higher risk of in-hospital gout flare than clopidogrel monotherapy (OR 1.57, 95% CI 1.07–2.31, P = 0.022). By enrolling only antithrombotic-naive patients, this study reduced confounding by prior antithrombotic exposure and helped clarify the association between aspirin-containing antiplatelet therapy and gout flare risk in this high-risk population.

Our findings point to two competing priorities: standardized antiplatelet regimens for stroke prevention and avoidance of aspirin-related gout flares. Aspirin-containing DAPT is increasingly used in acute ischemic stroke. In our cohort, the proportion of patients receiving DAPT increased substantially from 25.1% (2019–2021) to 50.8% (2024–2025). This rising trend was accompanied by improved standardization of DAPT prescription, with most indications now limited to mild stroke and ICAS, while off-standard indications dropped sharply from 39.1% to 15.0%. However, gout remains a barrier to maintaining this standard. In our cohort, aspirin discontinuation among patients with a flare stayed high (65.2% → 66.7% → 56.0%), while discontinuation among those without a flare fell steeply (47.8% → 28.6% → 11.9%). The gap between the two groups widened over time. For patients who meet guideline criteria for DAPT, stopping aspirin in the first days after symptom onset removes the early benefit of dual therapy. This real-world conflict suggests that clinicians should identify patients at high risk of gout flares and avoid unnecessary interruption of guideline-recommended antiplatelet therapy.

We next examined risk factors for flares in our cohort and whether they modified the effect of antiplatelet therapy. A related diagnostic concern is pseudogout (calcium pyrophosphate deposition disease, CPPD), which can mimic acute gout and lead to misclassification. CPPD is driven by elevated extracellular pyrophosphate concentrations within articular cartilage and triggers joint inflammation(Pascart et al. 2024). CPPD is more common in stroke patients with limited mobility or prolonged bed rest, and lesions predominantly affect paretic limbs or the atlantoaxial joint (Maki et al. 2008). In our cohort, baseline mRS ≥ 3 was independently associated with gout flare (OR 2.13, 95% CI 1.42–3.20, P < 0.001), overlapping with the CPPD-prone group. However, it did not appear to alter the antiplatelet–flare association: IPTW balancing yielded comparable mRS distributions between the aspirin and clopidogrel groups, and interaction analysis showed that functional status did not modify the association between antiplatelet therapy and flare risk. Thus, the elevated flare risk associated with aspirin appears unlikely to be substantially affected by CPPD misclassification. In practice, when severely disabled stroke patients receiving aspirin develop acute joint pain, CPPD should still be considered before diagnosing gout or adjusting antiplatelet therapy.

We further assessed whether baseline SUA levels and in-hospital ULT influenced gout flare risk or modified the aspirin–flare association. Our data showed that higher baseline SUA independently increased flare risk; each 1 mg/dL increment was associated with 20% higher odds of in-hospital gout flare. Short-term in-hospital benzbromarone did not reduce acute flares in this cohort. Neither baseline SUA nor ULT significantly modified the association between aspirin exposure and gout flares. Thus, aspirin-related gout activation does not appear to depend on baseline hyperuricemia and is unlikely to be attenuated by short-term uricosuric therapy. A possible explanation is that pre-existing hyperuricemia and deposited monosodium urate (MSU) crystals, together with changes in the internal milieu following acute ischemic stroke, may have primed the joints for inflammation via the NLRP3 inflammasome/IL-1β axis(Dalbeth et al. 2021); this primed state may not be rapidly reversible by lowering SUA. Future studies should explore whether targeting the NLRP3/IL-1β axis (e.g., with colchicine(Otani et al. 2016)) can mitigate this risk.

## 5 Limitations

Several limitations should be acknowledged. This was a retrospective, single-center observational study, and residual confounding cannot be excluded despite multivariable adjustment. Gout history and flare/recurrence relied on clinical documentation. For some patients, eligibility was determined after a gout flare through interview-based history, which may have introduced ascertainment and selection bias and limited the completeness of exposure assessment. The decision to interrupt DAPT after a gout flare may have been influenced by unmeasured clinical factors, so causal inference is not possible. We also lacked post-discharge follow-up and could not determine whether early DAPT interruption translated into a higher risk of recurrent ischemic stroke. These findings should be considered exploratory and require validation in prospective, multicenter studies with longer follow-up.

## 6 Conclusion

In AIS patients with a history of gout and hyperuricemia, aspirin-containing antiplatelet regimens (aspirin alone or dual therapy with clopidogrel) are associated with an increased risk of gout flare compared with clopidogrel monotherapy. This association was not modified by SUA level, mRS score, or ULT use. Elevated SUA and a baseline mRS score ≥ 3 were independent predictors of in-hospital gout flares. This study identifies an important real-world trade-off: gout flares commonly prompt aspirin discontinuation, which may compromise guideline-recommended DAPT. Clinicians should stratify gout flare risk prior to aspirin prescription and avoid unnecessary interruption of antiplatelet therapy.

## Data Availability

All data produced in the present study are available upon reasonable request to the authors

## Author Contributions

Aiwen Ma: Conceptualization, Data curation, Formal analysis, Investigation, Methodology, Project administration, Writing – original draft. Xiaolin Xu: Conceptualization, Supervision, Writing – review & editing. Yu Zhang: Data curation, Investigation, Writing – review & editing.

## Acknowledgments

None.

## Data Availability Statement

The datasets generated during and/or analysed during the current study are available from the corresponding author on reasonable request.

