## Supplementary Fig 1 for "Aspirin-Containing Antiplatelet Therapy and In-Hospital Gout Flares in Acute Ischemic Stroke Patients with Gout and Hyperuricemia: A Retrospective New-User Cohort Study"

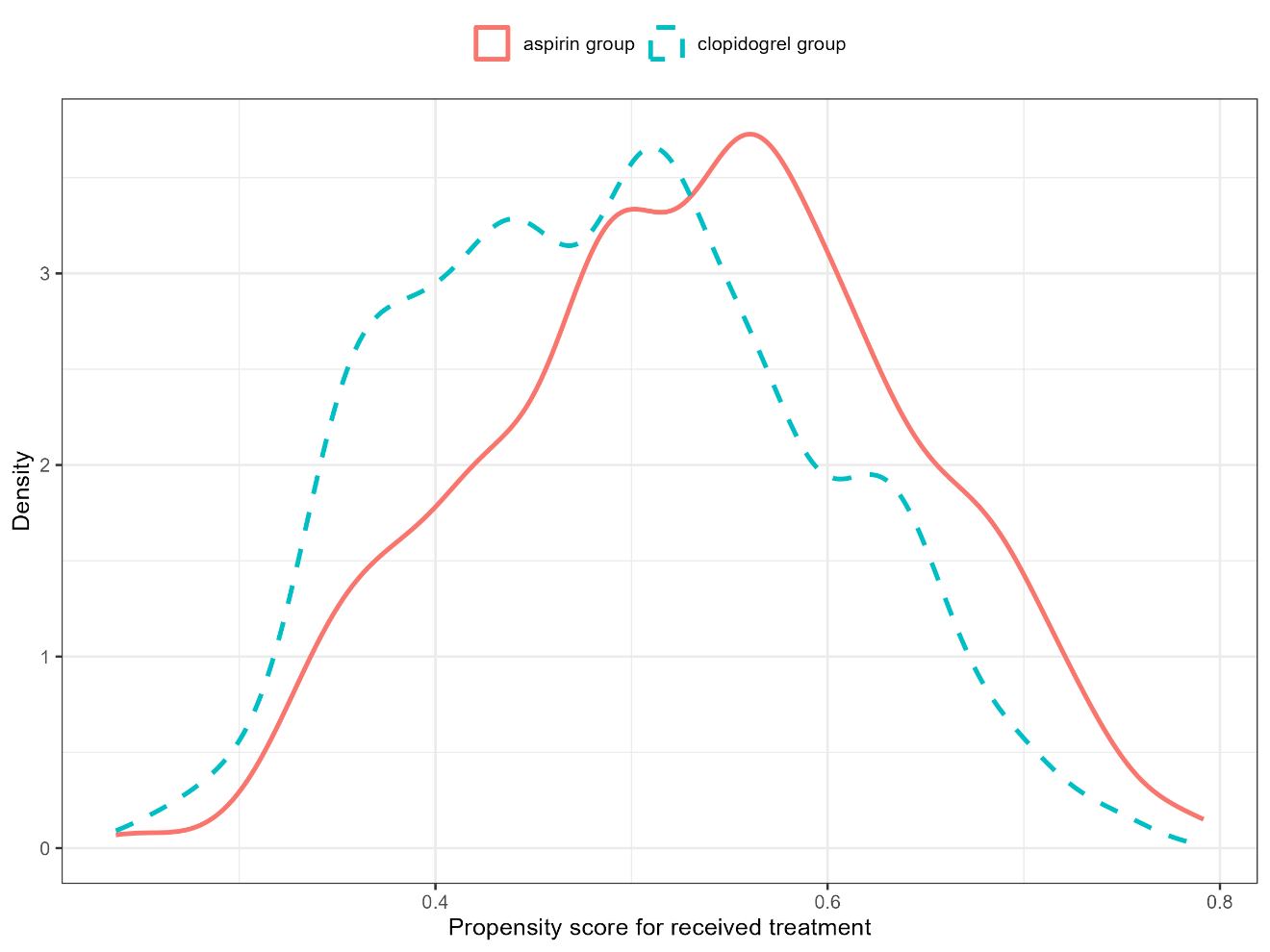

**Supplementary Fig 1.** Propensity score overlap diagnostic plot. The distributions of the propensity score for the received treatment are shown for each treatment group. Overlap across groups indicates that the positivity assumption is met.
