## Supplementary Fig 2 for "Aspirin-Containing Antiplatelet Therapy and In-Hospital Gout Flares in Acute Ischemic Stroke Patients with Gout and Hyperuricemia: A Retrospective New-User Cohort Study"

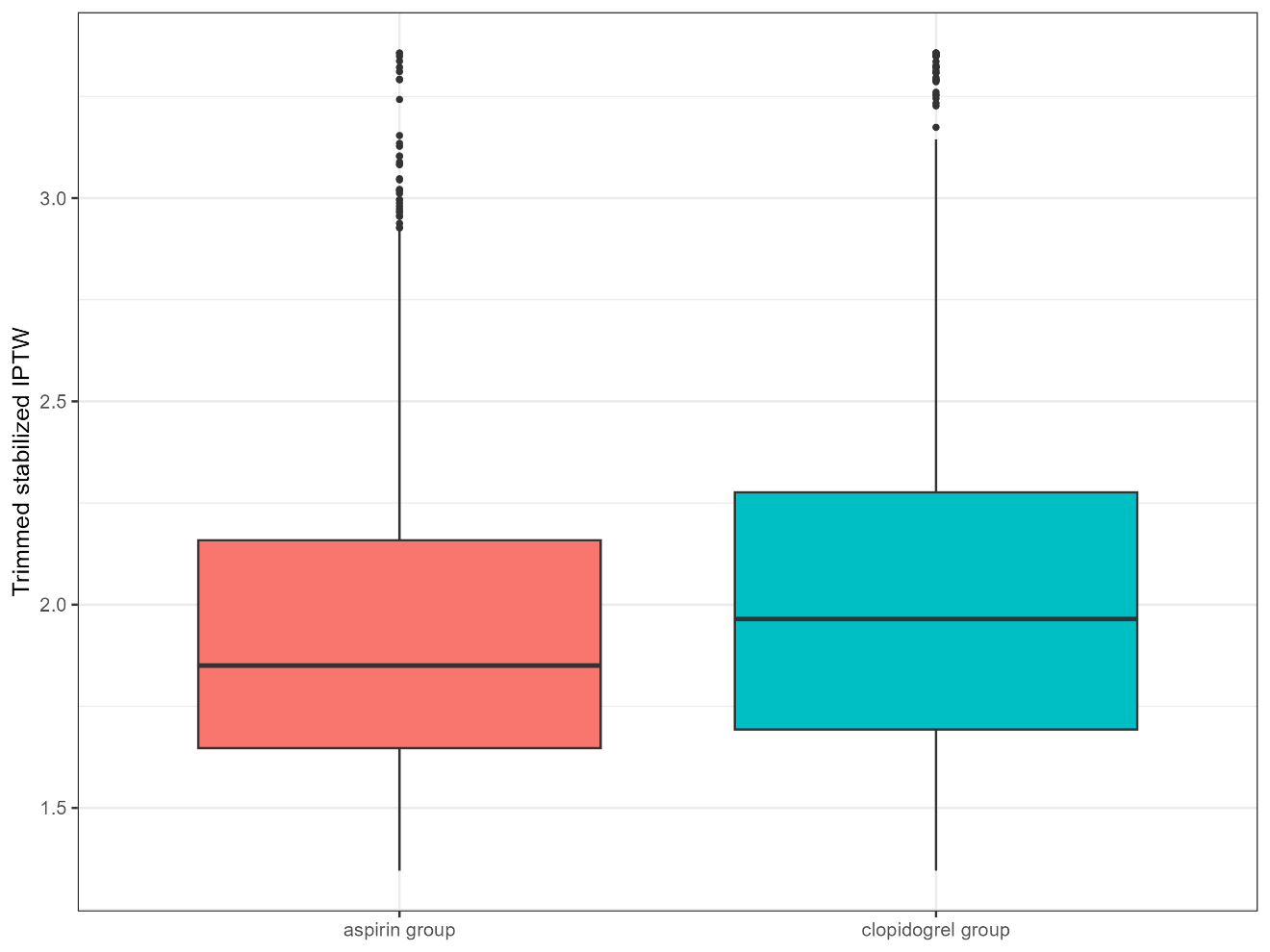

**Supplementary Fig 2.** Trimmed weight distribution. Weights were winsorized at the 1st and 99th percentiles. The density plot illustrates the distribution of the final weights applied in the main analysis.
